# Sex-Specific Effects of Adverse Childhood Experiences on Adolescent Brain Development: Insights from the ABCD Study

**DOI:** 10.1101/2025.06.08.25329159

**Authors:** Kangyi Peng, Shilat Haim-Nachum, Lawrence Maayan, Chen Zhang, Seonjoo Lee, Xi Zhu

## Abstract

**Background:** Adverse Childhood Experiences (ACEs) are established risk factors for physical and mental health outcomes, yet their associations with pediatric brain development remain underexplored, particularly regarding sex differences. Most studies treat ACEs as a single construct, overlooking the distinct effects of specific subtypes (e.g., sexual abuse, physical neglect) on brain structure and function. This study examines how ACE subtypes and their interaction with sex are linked to brain development in 9-10-year-olds.

**Methods:** Using data from the Adolescent Brain Cognitive Development (ABCD) Study (N≈12,000, ages 9–10), we assessed the links between emotional abuse, physical abuse, sexual abuse, emotional neglect, and physical neglect and structural brain measures in the hippocampus, amygdala, lateral orbital frontal cortex (lOFC), medial orbitofrontal cortex (mOFC), and rostral anterior cingulate cortex (rACC). Mixed-effects linear models tested ACE subtype, sex, and interaction effects, adjusting for multiple comparisons (Benjamini-Hochberg FDR correction).

**Results:** Boys reported higher exposure to emotional abuse (*p* =.01), emotional neglect (*p* = .037), and physical neglect (*p*< .001), while girls had greater exposure to sexual abuse (*p* < .001). Significant ACE-by-sex interactions emerged: sexual abuse was associated with smaller hippocampal volume in boys (*p* = .003) but showed no significant effect in girls (*p* = .433). Emotional abuse was linked to reduced lOFC volume in boys (*p* = 0.024), while in girls, it was associated with a marginal increase (*p* = .048). Physical neglect was associated with reduced hippocampal volume regardless of sex (pFDR = .05).

**Conclusion:** Boys may be particularly vulnerable to hippocampal and lOFC changes following sexual and emotional abuse, while physical neglect broadly impacts hippocampal development. These findings highlight sex-specific neurodevelopmental effects of ACE subtypes, emphasizing the need for tailored interventions and possible biomarkers for treatment and prevention of sequelae.

## Introduction

Adverse Childhood Experiences (ACEs) are well-established risk factors for poor physical and mental health outcomes (Felitti, 1998; Tzouvara et al., 2023). According to the 2016 National Survey of Children’s Health, nearly half (46.3%) of all children and 55.7% of adolescents in the United States have experienced at least one ACE (Bethell et al., 2017). Adolescence represents a sensitive period for brain maturation, during which the effects of early adversity may become more pronounced due to hormonal, social, and cognitive transitions (Backes & Bonnie, 2019). These neurodevelopmental changes can shape trajectories of emotional and cognitive functioning and increase vulnerability to psychiatric conditions in adulthood.

Extensive studies in adults have demonstrated that ACEs are linked to widespread structural and functional brain alterations, particularly in regions involved in emotion regulation, reward processing, and executive functions (Dillon et al., 2009; Herzog & Schmahl, 2018; Sheridan et al., 2017). Structural alterations have been observed in the hippocampus and amygdala, subcortical regions commonly associated with emotion regulation (Tottenham et al., 2010; Goff & Tottenham, 2015). These structures interact closely with prefrontal regions—notably the medial and lateral orbitofrontal cortices (mOFC, lOFC) and the rostral anterior cingulate cortex (rACC)—which are critical for affective evaluation, behavioral inhibition, and top-down regulation of emotional responses (Teicher et al., 2016; van Oort et al., 2025; Stinson et al., 2023). Likely related to these structural changes, ACEs have also been associated with altered functional connectivity in these emotion–regulation circuits. Resting-state studies show disrupted connectivity between the ventral striatum and prefrontal regions (Kamkar et al., 2017), as well as in the frontoparietal network (FPN)—a system essential for executive control and adaptive decision-making (McLaughlin et al., 2014). Notably, recent evidence from Yu et al. (2019) identified reduced connectivity in the cingulo-opercular network (CON), which supports sustained attention and cognitive stability, among adults with a history of childhood maltreatment. While findings in adults suggest robust brain alterations associated with ACEs, evidence in children and adolescents remains equivocal, possibly reflecting ongoing neurodevelopmental processes and methodological variability across studies.

Studies focusing on the association between youth brain and adverse experiences remain limited, often relying on relatively small sample sizes (typically ranging from 38 to 94 participants) and yielding mixed results. In pediatric populations, existing research suggests that maltreated children, regardless of whether they meet criteria for trauma-related disorders such as posttraumatic stress disorder (PTSD), tend to exhibit reduced total brain volume. This includes reductions in gray matter in the orbitofrontal cortex (OFC) and decreased area and white matter integrity in the corpus callosum (De Bellis et al., 2002; Carrion et al., 2001; Hanson et al., 2010; De Brito et al., 2013; for a review, see Bick & Nelson, 2016; Chahal et al., 2022). However, findings related to subcortical regions such as the hippocampus and amygdala have been less consistent. In contrast to robust evidence of reduced hippocampal and amygdala volumes in adults with childhood maltreatment histories, studies in children and adolescents often do not report such reductions (De Bellis et al., 2002; Carrion et al., 2001; De Brito et al., 2013). De Brito et al. (2023) suggested that the timing and type of adversity may differentially affect subcortical structures. For example, increased amygdala volume has been observed in children exposed to very early and severe institutional deprivation. These variable findings in pediatric samples, relative to the more consistent patterns observed in adults, underscore the complexity of neurodevelopment and highlight the need for large-scale datasets to better understand how early adversity shapes brain structure and function over time.

Large-scale neuroimaging datasets, such as the Adolescent Brain Cognitive Development (ABCD) Study, offer a unique opportunity to examine the impact of ACEs on brain structure and function. For example, hippocampal volume was significantly reduced in youth with greater severity of psychosis-like experiences (PLEs) and higher exposure to childhood stress (Damme et al., 2024). Greater ACE exposure has also been linked to reduced BOLD response in the right opercular region of the inferior frontal gyrus and bilateral pre-supplementary motor area – regions critical for inhibitory control (Stinson et al., 2024). Additionally, both ACEs and internalizing symptoms have been associated with reduced anti-correlation between the default mode network (DMN) and the dorsal attention network (DAN). ACEs were further linked to lower between-network connectivity between the auditory network and the cingulo-opercular network (CON), as well as heightened within-network connectivity of the frontoparietal network (FPN), highlighting widespread alterations in functional connectivity patterns (Albertina et al., 2024).

Despite growing research on ACE-related brain changes, key gaps remain in the analytical approaches used. First, most current ABCD studies assess ACEs using cumulative risk scores, categorizing exposure as none (0), moderate (1–3), or high (4+), or by dichotomizing risk based on threat vs. deprivation (e.g., Damme et al., 2024; Stinson et al., 2024; Xiao et al., 2024). While useful, this approach may obscure the distinct effects of individual ACE subtypes (e.g., sexual abuse, physical neglect) on brain development. Previous research has suggested that specific ACE subtypes – such as neglect, sexual abuse, and emotional abuse –exert unique effects on brain development (Teicher & Samson, 2016). For instance, childhood neglect, including both physical neglect and emotional neglect, has been associated with increased ventral diencephalon volume, greater vulnerability to reward-related disorders, and more diffuse white matter organization in the prefrontal cortex and connected regions (Teicher & Samson, 2016; Hanson et al., 2013). Sexual abuse has been linked to hippocampal volume changes (Andersen et al., 2008); emotional abuse and neglect have been associated with increased amygdala volume (Pechtel et al., 2014); and physical abuse has been related to smaller OFC volume (Hanson et al., 2010).

These findings highlight the importance of considering ACE subtypes individually when examining neurodevelopmental consequences. Assigning equal weight to all ACE subtypes in cumulative scoring may overlook the differential neurodevelopmental impact of specific forms of adversity and limit possible interpretations for cognitive and emotional functions depending on different neural mechanisms.

Second, few large-scale studies have systematically examined how specific ACE subtypes affect brain biomarkers. For example, a large-scale study using the European YOUth cohort found that household substance abuse was linked to greater cortical surface area in several frontal regions (including the left and right superior frontal gyrus, left pars triangularis, left rostral middle frontal gyrus, and right caudal anterior cingulate gyrus). In contrast, household exposure to violence was associated with lower fractional anisotropy in the left and right cingulum bundle hippocampal regions (Buimer et al., 2022). However, due to data limitations, that study did not examine certain common subtypes (e.g., sexual, emotional, physical abuse) or broader dichotomous classifications (abuse vs. neglect), which are commonly used in psychological and clinical research to define childhood maltreatment (Haim-Nachum et al., 2024). Incorporating these more nuanced subtypes with a large sample size is essential to advancing our understanding of neurodevelopmental risk.

Third, many current studies are underpowered and, for this and other methodological reasons, often fail to adequately examine the specific impact of sex differences on the relationship between ACEs and brain structure. However, the limited evidence that does exist suggests sex differences in the brain’s developmental response to early adversity, though findings are more mixed in pediatric samples than in adults. In adult samples, hippocampal volume, when measured in males, appears to be influenced more by neglect in early childhood, whereas in females, it is more strongly affected by abuse during preadolescence (between ages 10 and 11) (Teicher et al., 2018). Several studies have also shown heightened vulnerability among adult males to the neurobiological consequences of early adversity. For example, neglect and emotional abuse were linked to reduced hippocampal volume in males but not in females (Samplin et al., 2013; Frodl et al., 2010; Everaerd et al., 2012). Similarly, trauma exposure has been more strongly associated with hippocampal reductions in males than in females (Karl et al., 2006). In pediatric samples, however, findings are less consistent. Trauma-exposed boys have shown stronger rsFC within the DMN than girls (Wang et al., 2021). Adolescents with histories of childhood abuse and neglect demonstrate increased within-salience network (SN) connectivity, with boys showing neglect-related increases in within-DMN connectivity not seen in girls (Rakesh et al., 2023). Nonetheless, large-scale studies that explicitly examine sex differences in structural and functional brain features following childhood trauma remain scarce. Even within the same cohort, results can vary: For instance, Rakesh et al. (2023) reported no amygdala or hippocampal volumetric differences in adolescents exposed to childhood abuse and neglect, except for increased amygdala volume associated with greater neglect among girls only.

To address these limitations, the current study leverages the ABCD dataset to examine individual ACE subtypes—emotional abuse, physical abuse, sexual abuse, emotional neglect, and physical neglect—and their distinct associations with childhood brain structure and function. We focused on these five ACEs subtypes, as these forms of maltreatment are widely recognized in both research and clinical practice as distinct categories of child abuse and neglect. They are commonly used in psychological and neurodevelopmental research due to their well-defined impact on a child’s emotional, cognitive, and physical development (for a similar approach, see Haim-Nachum et al., 2024; Grummitt et al., 2022; Massullo et al., 2023). Additionally, we investigate whether sex moderates these associations, given growing evidence for sex-specific neurodevelopmental responses to adversity. We hypothesize that (1) sex differences will emerge in exposure to specific ACE subtypes, (2) sex will moderate the relationship between ACE subtypes and brain outcomes, and (3) greater exposure to ACE subtypes will be associated with reduced volumes or rsFC in brain regions involved in emotional regulation and reward processing (e.g., hippocampus, amygdala, lOFC, mOFC, rACC) in both boys and girls.

## 1. Methods

### 1.1. Participants and design overview

The current study utilized baseline data from the ABCD Study®, a longitudinal cohort of 11,889 children recruited between 2016 and 2018 at ages 9–10 years from 21 sites across the United States. Participants were recruited through a stratified probability sampling approach to ensure that the sample was demographically representative of the U.S. population based on sex, race/ethnicity, socioeconomic status, and urbanicity. Study procedures were approved by the centralized Institutional Review Board at the University of California, San Diego, and informed parental consent and youth assent were obtained at each visit. At baseline, participants and their caregivers attended one to two sessions, during which they completed a comprehensive battery of questionnaires and underwent structural and functional magnetic resonance imaging (MRI) and cognitive assessments. For the present study, only baseline neuroimaging and clinical data were analyzed to examine the cross-sectional associations between ACEs, sex, and brain structure and function.

### 1.2. Adverse Childhood Experiences

The current study measured five subtypes of ACEs: Emotional abuse, physical abuse, sexual abuse, emotional neglect, and physical neglect. Items were adapted from previous ACE measures used in Hill et al. (2025) and Damme et al. (2024) (see Supplementary Materials for details). Emotional abuse, physical abuse, and sexual abuse were answered by parents. Emotional neglect and physical neglect were answered by the youth.

#### Emotional Abuse

This construct was assessed using two binary items (0 = No, 1 = Yes) adapted from Damme et al. (2024): (1) “A non-family member threatened to kill your child”

(*ksads_ptsd_raw_764_p*) and (2) “A family member threatened to kill your child” (*ksads_ptsd_raw_765_p*). An emotional abuse score of 1 was assigned if either item was endorsed.

#### Physical Abuse

Two binary items were used (0 = No, 1 = Yes), adapted from Damme et al. (2024) and Hill et al. (2025): (1) “Shot, stabbed, or beaten brutally by a grown-up in the home” (*ksads_ptsd_raw_762_p*), and (2) “Beaten to the point of having bruises by a grown-up in the home” (*ksads_ptsd_raw_763_p*). A physical abuse score of 1 was assigned if either item was endorsed.

#### Sexual Abuse

This was measured using two binary items (0 = No, 1 = Yes), adapted from Damme et al. (2024) and Hill et al. (2025): (1) “Grown-up in the home touched your child in their privates, had your child touch their privates, or did other sexual things to your child” (*ksads_ptsd_raw_767_p*), and (2) “An adult outside your family touched your child in their privates, had your child touch their privates, or did other sexual things to your child” (*ksads_ptsd_raw_768_p*). A sexual abuse score of 1 was assigned if either item was endorsed.

#### Emotional Neglect

Ten items were used on a 3-point scale (1 = Not like him/her, 2 = Somewhat like him/her, 3 = A lot like him/her), along with one binary item used to assess the presence of an additional caregiver. These items were adapted from Hill et al. (2025). Sample items include: “Parent or caregiver makes me feel better after talking over my worries with them” (*crpbi_parent1_y*) and “a parent or a caregiver smiles at me very often” (*crpbi_parent2_y*). If a participant had additional caregivers, the mean was calculated across ten scaled items (five per caregiver); if only one caregiver was present, the mean was computed across five scaled items. Participants with a mean score below 2 were coded as experiencing emotional neglect (score = 1).

#### Physical Neglect

Five items with a 5-point numeric scale, adapted from Damme et al. (2024). For example, “How often do your parents/guardians know where you are?” (parent_monitor_q1_y), and “How often do your parents know who you are with when you are not at school and away from home?” (parent_monitor_q2_y) (1 = Never; 2 = Almost Never; 3 = Sometimes; 4 = Often; 5 = Always or Almost Always). A mean score was computed across all five items. Participants with a mean score below 3 were coded as experiencing physical neglect (score = 1).

### 1.3. Imaging processing

Structural MRI: Participants completed a high-resolution T1-weighted structural MRI scan (1-mm isotropic voxels). Structural MRI data were processed using FreeSurfer version 7.0 (http://surfer.nmr.mgh.harvard.edu/) according to standard processing pipelines. Processing included the removal of non-brain tissue and the segmentation of gray and white matter structures. Quality control for the structural images included visual inspection of T1 images and FreeSurfer outputs for quality, conducted by the ABCD team. Subjects whose scans failed inspection or were identified as extreme outliers by Rosner’s test were excluded. Brain structural volumes were defined by the Desikan-Killiany atlas. For each region of interest (ROI), total volume was calculated by summing the left and right hemisphere volumes. ETIV represents the sum of total brain tissue and cerebrospinal fluid (CSF) within the cranial cavity and serves as a scaling factor for individual differences in brain size. Following standard neuroimaging protocols, ETIV was included as a covariate in volumetric analyses to account for variations in head size across participants (ABCD Study Consortium, 2023). In line with the current literature mentioned above, the current study examined volume measures for the hippocampus, amygdala, lateral orbitofrontal cortex (lOFC), medial orbitofrontal cortex (mOFC), and rostral anterior cingulate cortex (rACC).

Functional MRI: Participants also completed a resting-state functional MRI (rs-fMRI) scan, collected using multi-band echo-planar imaging sequences (TR = 800 ms, TE = 40, voxel size = 2.4 mm isotropic, slices = 60). Resting-state fMRI data were preprocessed using the ABCD-HCP BIDS pipeline, which includes motion correction, slice timing correction, co-registration to structural images, spatial normalization to MNI space, and smoothing with a 6-mm full-width at half-maximum (FWHM) Gaussian kernel. For resting-state models, only participants with high-quality resting-state fMRI data (i.e., those with imgincl_rsfmri_include = 1) were included in the analyses. We examined within-network connectivity in the SN, DMN, FPN, and CON, as well as between-network connectivity among SN-DMN, SN-FPN, and DMN-FPN, based on the Gordon atlas.

### 1.4 Statistical Analysis

Data preprocessing began by merging baseline datasets, identifying and removing duplicate participant entries (n = 44), and cleaning non-informative or missing values. Age was converted from months to years for interpretability. Values indicating non-responses or inapplicability (e.g., 888 for race_ethnicity and siblings_twins; 777, 888, and 999 for income) were recorded as missing values (NA) in R. This ensured that only meaningful data contributed to each analysis. All statistical analyses were conducted using R and Python. Demographic analyses (e.g., chi-square tests, descriptive statistics) were performed in R, while imaging data processing and statistical modeling were conducted in Python. To maximize the inclusion of available imaging data, missing values were handled on a per-model basis (i.e., participants were excluded only if missing data for the specific analysis), avoiding listwise deletion.

Chi-square tests were used to examine sex differences in exposure to each ACE subtype: emotional abuse, physical abuse, sexual abuse, emotional neglect, and physical neglect. These analyses included all participants with complete data on sex and the relevant ACE variables.

For analyses involving brain structural data, only participants with high-quality MRI scans imgincl_t1w_include = 1) were included. Estimated total intracranial volume (ETIV) was standardized using z-scores (ETIV_z) and included as a covariate in all models involving brain volume to account for individual differences in head size.

To test whether sex moderates the association between the five ACE subtypes and brain structure and function, mixed-effect linear models were performed for each brain region of interest (hippocampus, amygdala, lOFC, mOFC, and rACC). Each model included the interaction between five ACE subtypes and sex when controlling for ETIV, and site as a random effect. False Discovery Rate corrections (FDR) were performed to adjust the False Positive rate.

To test whether sex moderated the relationship between ACEs and brain volume, mixed-effects linear regression models were performed for each brain region of interest (ROI): hippocampus, amygdala, lOFC, mOFC, and rACC. Each model included one ACE subtype, sex, the ACE × sex interaction, and ETIV_z as a covariate. The site was modeled as a random effect. FDR correction was applied to control for multiple comparisons across the 25 interaction models (5 ACE subtypes × 5 ROIs). An example model specification is as follows: Hippocampus_total ∼ ace1_emotional_abuse + sex + ace1_emotional_abuse * sex + ETIV_z, random effect = site.

An additional set of mixed-effects models was conducted to examine the main effects of ACE subtypes on brain volume. In these models, the ACE subtype was included as the primary fixed effect, with sex and ETIV_z included as covariates. The site was modeled as a random effect. The same FDR correction was applied across the 25 comparisons (5 ACE subtypes × 5 brain regions). An example model specification is as follows: Hippocampus_total ∼ ace1_emotional_abuse + sex + ETIV_z, random effect = site.

All mixed-effects models—both main effect and interaction models—were accounted for multiple comparisons, FDR correction was applied across a total of 60 comparisons: 5 structural regions of interest (hippocampus, amygdala, lOFC, mOFC, and rACC), 7 resting-state connectivity measures (within SN, within DMN, within FPN, within CON, between SN-DMN, between SN-FPN, and between DMN-FPN), and 5 ACE subtypes (emotional abuse, physical abuse, sexual abuse, emotional neglect, and physical neglect).

## 2. Results

### 2.1. Participants

A total of 44 duplicate participants from site 22 were identified and removed, resulting in 18 unique participants retained from site 22. Although the ABCD study officially includes 21 data collection sites, some participants visited multiple locations (e.g., for different types of assessments), resulting in additional site entries. To retain as much usable neuroimaging data as possible, we kept the 18 non-duplicated participants from site 22 in the sample. After excluding participants with missing sex data, the final sample included 11,866 individuals (47.9% assigned female at birth) with a mean age of 9.91 years (*SD* = 0.62), prior to T1 data quality control (see Table 1). This sample was used for analyses involving demographic and clinical characteristics. Participants with non-informative responses (e.g., coded as 777, 888, or 999) were excluded from analyses related to race/ethnicity, sibling/twin status, and household income. Following T1 data quality control, 11,218 individuals remained for analyses involving brain structural data.

**Table 1.** Demographic and clinical characteristics by sex at baseline, N=11,866.

| **Characteristic** | **Male**<br>N = 6,185<sup>1</sup> | **Female**<br>N = 5,681<sup>1</sup> | ***p*-value<sup>2</sup>** |
| --- | --- | --- | --- |
| **Age (years)** | 9.92 (0.62) | 9.90 (0.62) | 0.023* |
| **Race/Ethnicity** |  |  | 0.12 |
| White | 3,274 (53%) | 2,899 (51%) |  |
| Black | 886 (14%) | 891 (16%) |  |
| Hispanic | 1,248 (20%) | 1,157 (20%) |  |
| Asian | 121 (2.0%) | 130 (2.3%) |  |
| Other | 646 (10%) | 599 (11%) |  |
| Missing | 10 | 5 |  |
| **Siblings/Twins** |  |  | 0.14 |
| Only child | 4,304 (70%) | 3,841 (68%) |  |
| Has siblings (not twins) | 803 (13%) | 786 (14%) |  |
| Dizygotic twin | 1,064 (17%) | 1,038 (18%) |  |
| Monozygotic twin | 14 (0.2%) | 16 (0.3%) |  |
| **Household Income** |  |  | 0.4 |
| <\$5,000 | 219 (3.9%) | 197 (3.8%) | |
| \$5,000–11,999 | 211 (3.7%) | 211 (4.0%) | |
| \$12,000–15,999 | 140 (2.5%) | 133 (2.6%) | |
| \$16,000–24,999 | 287 (5.1%) | 235 (4.5%) | |
| \$25,000–34,999 | 313 (5.5%) | 340 (6.5%) | |
| \$35,000–49,999 | 478 (8.5%) | 456 (8.8%) | |
| \$50,000–74,999 | 797 (14%) | 701 (13%) | |
| \$75,000–99,999 | 804 (14%) | 767 (15%) | |
| \$100,000–199,999 | 1,745 (31%) | 1,569 (30%) | |
| ≥\$200,000 | 647 (11%) | 602 (12%) | |
| Missing | 544 | 470 |  |
| **ACE Exposure** |  |  |  |
| **Emotional Abuse** | 75 (1.2%) | 41 (0.7%) | 0.010* |
| Missing | 140 | 159 |  |
| **Physical Abuse** | 55 (0.9%) | 40 (0.7%) | 0.317 |
| Missing | 135 | 154 |  |
| **Sexual Abuse** | 31 (0.5%) | 65 (1.2%) | <0.001*** |
| Missing | 135 | 154 |  |
| **Emotional Neglect** | 59 (1.0%) | 34 (0.6%) | 0.037* |
| Missing | 14 | 15 |  |
| **Physical Neglect** | 198 (3.2%) | 92 (1.6%) | <0.001*** |
| Missing | 10 | 12 |  |

<sup>1</sup>Mean (SD); n (%)
<sup>2</sup>Welch Two Sample t-test; Pearson's Chi-squared test; \*p<0.05; \*\*p<0.01; \*\*\*p<0.001

### 2.2. Sex difference in exposure to five ACE subtypes

Significant associations were observed between sex and emotional abuse, sexual abuse, emotional neglect, and physical neglect. Boys were more likely than girls to be exposed to emotional abuse, *χ²*(1, *N* = 11,844) = 6.72, *p* = .010; emotional neglect, *χ²*(1, *N* = 11,844) = 4.36, *p* = .037; and physical neglect, *χ²*(1, *N* = 11,844) = 30.37, *p* < .001. In contrast, girls were more likely than boys to be exposed to sexual abuse, *χ²*(1, *N* = 11,844) = 14.67, *p* < .001. No significant association was found between sex and physical abuse, *χ²*(1, *N* = 11,844) = 1.00, *p* = .317.

### 2.3 Interaction between ACE subtypes and sex on brain structure

For hippocampal total volume, a significant interaction between sexual abuse and sex was observed, *β* = 418.752, *SE* = 139.704, 95% CI [144.938, 692.567], *p* = .003. This effect remained significant after FDR correction (pFDR = .038). To further explore this interaction, follow-up mixed-effects models were conducted separately by sex. Among boys, greater exposure to childhood sexual abuse was significantly associated with lower hippocampal volume, *B* = −343.63, *SE* = 116.85, *z* = −2.94, *p* = .003. In contrast, the association was not significant in girls, *B* = 61.11, *SE* = 77.95, *z* = 0.78, *p* = .433 (see Figure 1).

**Figure 1.**
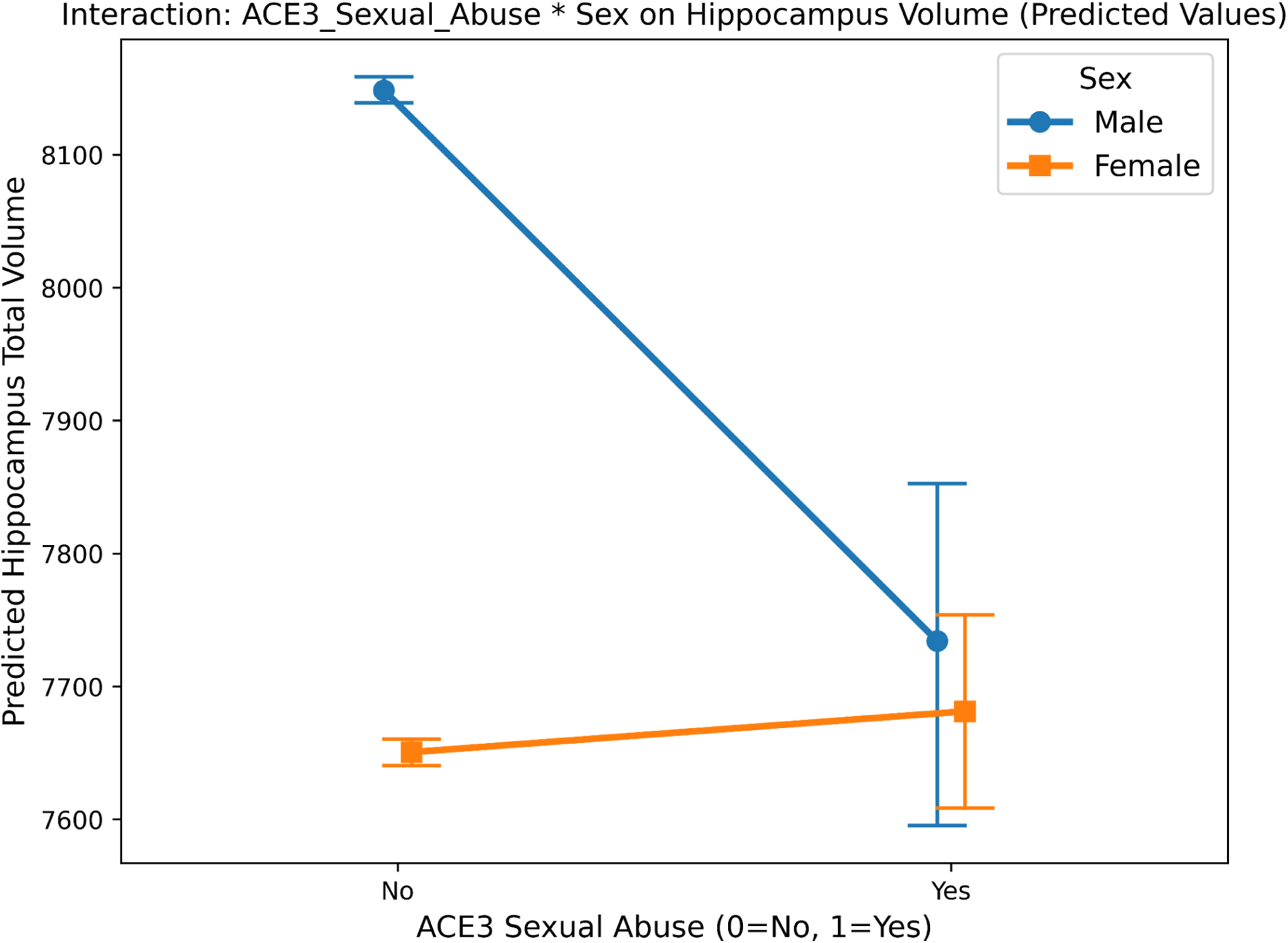
Interaction Between Sexual Abuse and Sex on Hippocampus Predicted Volume

For lOFC volume, a significant interaction between emotional abuse and sex was also found, *β* = 997.21, *SE* = 338.663, 95% CI [333.443, 1660.978], *p* = .003. This interaction remained significant after FDR correction (pFDR = .038). Follow-up mixed-effects models showed that in boys, higher exposure to emotional abuse was significantly associated with reduced IOFC volume, *B* = −456.64, *SE* = 202.83, *z* = −2.25, *p* = .024. Among girls, the association was marginally significant, *B* = 531.54, *SE* = 269.24, *z* = 1.97, *p* = .048, suggesting that emotional abuse may be associated with increased IOFC volume in females (see Figure 2).

**Figure 2.**
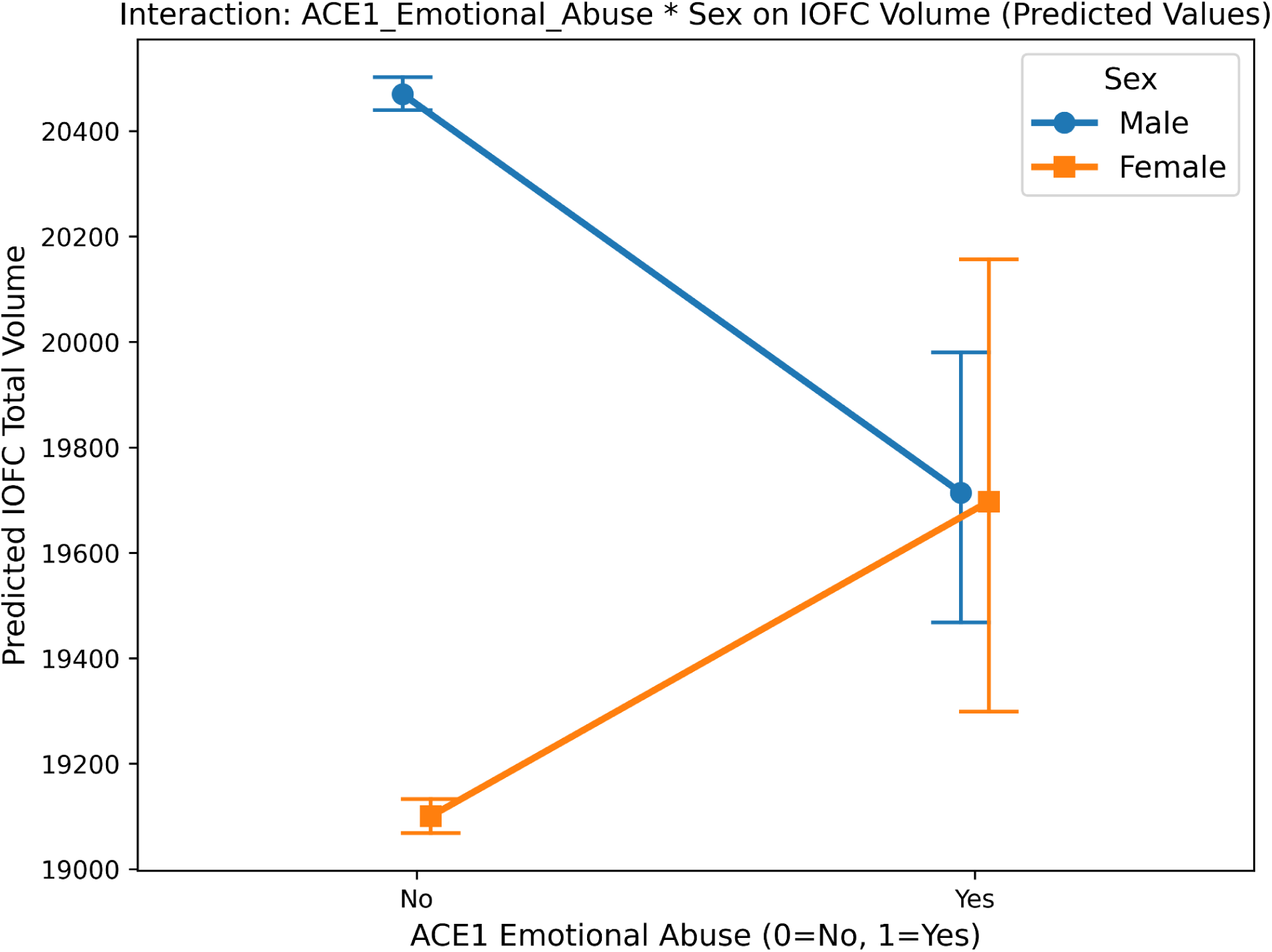
Interaction Between Emotional Abuse and Sex on IOFC Predicted Volume

These findings suggest that the effects of childhood sexual and emotional abuse on brain volume differ by sex, with certain adverse experiences having a stronger structural impact in one sex than the other. No significant FDR-corrected interaction effects were observed for the amygdala, rACC, mOFC, or rACC volumes.

### 2.4 Main effect of ACE subtypes on brain structure

For total hippocampal volume, a significant main effect of physical neglect was observed while controlling for sex, *β* = −118.20, *SE* = 38.96, 95% CI [−194.55, −41.84], *p* = .002. This effect remained marginally significant after FDR correction (pFDR = .050). These results suggest that greater exposure to physical neglect is associated with reduced hippocampal volume, regardless of sex; however, this finding warrants further investigation.

### 2.5 fMRI rsFC

No significant associations between the five ACE subtypes and functional brain connectivity were found.

## Discussion

This study examined whether subtypes of ACEs are differentially associated with brain structure in early adolescence and whether these associations vary by sex. The findings revealed that boys in the sample experienced more emotional abuse, emotional neglect, and physical neglect than girls, whereas girls experienced more sexual abuse. Importantly, we found that sexual abuse, emotional abuse, and physical neglect were linked to structural differences in the hippocampus and OFC. Several of these associations differed by sex: sexual abuse was associated with smaller hippocampal volume in boys but not in girls, while emotional abuse was related to reduced lOFC volume in boys but a marginal increase in girls. In contrast, physical neglect was associated with reduced hippocampal volume across both sexes. Taken together, these results highlight both the sex differences in exposure to ACE subtypes as well as sex-specific patterns in the associations between ACEs and brain structural volumes among adolescents.

### Male-Female differences in ACE prevalence

Our findings show that girls experienced higher rates of sexual abuse, while boys experienced higher emotional abuse as well as emotional and physical neglect. Prior research indicates that girls are more likely than boys to experience sexual abuse, with global estimates suggesting that 1 in 5 women and 1 in 7 men report such experiences during childhood (World Health Organization, 2024). Stoltenborgh et al. (2013) reported no significant gender differences in physical or emotional neglect in a meta-analysis; however, among Swedish adolescents, boys reported similar prevalence rates of emotional neglect compared to girls but reported a higher degree of emotional abuse (Hagborg, Tidefors, & Fahlke, 2017). In contrast, among Chinese adolescents living in rural regions, boys were more likely to experience physical abuse, while girls were more susceptible to emotional neglect (Wan et al., 2021). These inconsistent findings may be due to differences in ACE measurement, culture, or social norms.

### Male-female differences in the relationship between sexual abuse and hippocampal volume

We observed a dissociation between higher rates of sexual abuse exposure in girls and greater hippocampal volume reduction in boys. This might indicate greater vulnerability of the male hippocampus compared to the female hippocampus. While greater hippocampal vulnerability in maltreated males compared to females has not been reported in pediatric samples, it has been observed in several studies using adult samples (Samplin et al., 2013; Frodl et al., 2010; Everaerd et al., 2012; Karl et al., 2006). Prior research on childhood sexual abuse has predominantly focused on female survivors, with relatively little attention paid to the neurodevelopmental consequences of such abuse among males, particularly in childhood and adolescence (Gartner, 2018). Existing clinical studies involving adult male survivors have examined treatment outcomes (e.g., Gallo-Silver et al., 2014; Sivagurunathan et al., 2019), but have not included neuroimaging, leaving a critical gap in understanding sex-specific brain alterations during sensitive developmental windows. To our knowledge, the present study is the first to demonstrate that boys exposed to sexual abuse exhibit greater reductions in hippocampal volume in a large, population-based pediatric sample, despite girls reporting a significantly higher rate of sexual abuse exposure compared to boys.

The non-significant finding in girls’ hippocampal volume is consistent with Sussman et al. (2022), who explored the impact of sexual abuse-related PTSD occurring within the previous six months in female adolescents. However, it contrasts with findings from Demir et al. (2023), who reported larger hippocampal and amygdala volumes in female adolescents exposed to sexual abuse. This discrepancy may be driven by differences in clinical status (presence or absence of PTSD) and the timing of sexual abuse exposure. Although several studies using adult samples have consistently found reduced hippocampal volume in females with a history of sexual abuse (Riem et al., 2015; Stein et al., 1997), more research using pediatric samples is needed.

The non-significant finding in girls’ hippocampal volume may be explained by sex-specific sensitive periods for hippocampal vulnerability. For example, Andersen et al. (2008) reported that sexual abuse occurring between ages 3–5 and 11–13 was associated with reduced hippocampal volume in young adult women, while Humphreys et al. (2019) found that early life stress experienced through age 5 predicted alterations in adolescent hippocampal structure. In the present study, while neuroimaging was conducted at ages 9–10, the specific onset and duration of maltreatment were not reported. Thus, the abuse may have occurred before or after these sensitive periods, limiting our ability to conclude age-specific neurodevelopmental risk. It is also possible that protective factors—such as higher rates of trauma disclosure or closer social connections—may buffer girls against the neurodevelopmental effects of adverse experiences (Sivagurunathan et al., 2018). Finally, the neural consequences of sexual abuse in girls may manifest in brain regions other than the hippocampus, or specific hippocampal subregions not captured in our analysis. For example, childhood sexual abuse exposure has been linked to cortical thinning in the somatosensory cortex (Heim et al., 2013), alterations in the corpus callosum during late childhood (Andersen et al., 2008), and reduced grey matter in occipital and fusiform regions involved in facial recognition (Tomoda et al., 2009). The large-scale neuroimaging data available in ABCD may enable future studies to more comprehensively explore these sex-specific effects across more nuanced brain region parcellation and possibly neuroplastic adaptive responses in specific brain regions of different sexes.

### Male-Female Differences in the Relationship Between Emotional Abuse and OFC Volume

Emotional abuse was associated with reduced lOFC volume in boys, but a marginal increase in girls. These findings contribute to the growing view that emotional abuse can have lasting neurobiological consequences, comparable to more commonly studied forms of maltreatment (Dye, 2020; Brassard et al., 2000). While some previous studies have combined emotional abuse and neglect into a single construct (e.g., Pechtel et al., 2014), our results showed no significant effects of emotional neglect, suggesting that emotional abuse may have distinct associations with brain development.

Given that boys in our sample were more likely to be exposed to emotional abuse and neglect, the observed reduction in lOFC volume may not be solely driven by higher exposure but could instead reflect a sex-specific neural response. This aligns with prior findings showing that childhood emotional abuse was associated with reduced hippocampal volume in adult males but not females (Samplin et al., 2013).

Preclinical studies have indicated that early life stress affects hippocampal and prefrontal regions differently depending on developmental timing (Andersen & Teicher, 2004, 2008). Thus, future research using pediatric samples should explore whether girls’ lOFC may be more resilient to emotional abuse during specific developmental windows.

While our study did not find significant effects of emotional abuse on other brain regions, prior research in adult samples has linked emotional abuse to cortical thinning in the precuneus and anterior/posterior cingulate cortex—regions involved in self-referential processing and part of the default mode network (Heim et al., 2013). In terms of altered emotional processing, an EEG study by Liu et al. (2021) found that young men with a history of emotional abuse recognized negative facial expressions more quickly and with fewer cognitive resources, possibly reflecting heightened sensitivity to threat-related cues. Future research could explore how facial recognition processes interact with the lOFC in pediatric samples, and whether such alterations are protective or maladaptive in the long run.

### Main Effects of ACEs on Brain Structure Physical neglect and brain

In the current study, physical neglect was associated with reduced hippocampal volume across sexes. Limited previous research has examined the unique effects of physical neglect independent of other maltreatment types. For example, Frodl et al. (2010) found smaller hippocampal white matter in male adults who experienced childhood physical neglect but not in female adults. Our results extend this work by identifying gray matter volume reductions in a pediatric sample, regardless of sex. These differences may reflect developmental stage, measurement modality (white vs. gray matter), or varying patterns of brain maturation and compensation across sexes. Together, these findings support the hippocampus as a sensitive region affected by early physical neglect, though the precise nature and timing of these alterations may differ by sex and tissue type.

No significant associations were found with other predefined ROIs. Prior work has linked physical neglect to the amygdala and ACC, although findings vary based on measurement approaches, sample characteristics, and developmental timing. For example, broader reviews suggest that neglect is particularly associated with altered development in ACC. Teicher and Samson (2016) identified reduced ACC volume as one of the most consistent findings across studies of childhood maltreatment, including neglect. Future research using pediatric samples might benefit from expanding the ROI to the full ACC, which has been shown to be especially susceptible to neglect in adult males (Teicher et al., 2004), and expanding to explore if physical neglect alone can drive ACC alternation and when these changes begin to emerge.

The non-significant association between the amygdala and physical abuse in the current study corresponds to previous literature suggesting that amygdala changes may be time-sensitive and non-linear (Teicher & Samson, 2016). Specifically, increased amygdala volume has been observed during childhood in response to early neglect (Mehta et al., 2009; Tottenham et al., 2010; Lupien et al., 2011; Pechtel et al., 2014), while reductions have been found in older adolescents or adults (Hanson et al., 2015; Gerritsen et al., 2015). Given our participants were scanned at ages 9–10, without information on the onset and duration of maltreatment, it is possible they were in a transitional phase during which amygdala alterations had not yet emerged or had already normalized. The timing, chronicity, and type of neglect may play a crucial role in shaping amygdala development, and without precise temporal information, detecting such changes may be challenging.

Variability in findings may also reflect differences in measurement approaches. In this study, physical neglect primarily reflected lack of parental supervision—e.g., limited parental monitoring of whereabouts or daily interactions—rather than deprivation of basic survival needs. In contrast, many prior studies (e.g., those using the CTQ) have operationalized physical neglect as failure to provide food, shelter, medical care, or physical safety. Given that the amygdala is central to processing threat and fear, neglect involving unmet survival needs may be more likely to affect its development. Thus, future studies should further delineate different dimensions of physical neglect, such as lack of supervision versus deprivation of basic care, and examine their potentially distinct neural correlates. Clarifying these distinctions may help improve both research validity and clinical assessment, particularly when adapting definitions across cultural and legal systems (Zeanah & Humphreys, 2018).

### Interpretation of Null Findings of Functional Connectivity

We examined functional connectivity at a single developmental time point and did not find significant associations with the five ACE subtypes. In contrast, Chahal et al. (2022) employed a longitudinal design and found that maltreated youth exhibited steeper age-related declines in resting-state connectivity within and between the SN, DMN, and FPN. Rakesh et al. (2023) found increased SN connectivity following abuse or neglect, with neglect-related increases in DMN connectivity in male adolescents only, with two scans at 16 years old and 19 years old. Suzuki et al. (2014) detected a greater response of the amygdala to negative facial expressions in children experiencing greater stress. Future research leveraging longitudinal data could explore whether functional connectivity differences associated with ACEs emerge or evolve over time.

Previous studies using adult samples have explored broader network connectivity. For example, Chiasson et al. (2022) reported reduced functional connectivity in the striatal-thalamic circuits of the SN and weaker connectivity between the bilateral amygdalae and medial prefrontal cortex (mPFC) in adult men with histories of childhood sexual abuse compared to non-abused peers. Task-based studies have similarly shown that men with childhood sexual abuse and PTSD exhibit reduced activation in the cerebellum and fusiform gyrus (Chiasson et al., 2021), as well as greater mPFC connectivity within the default mode network (Chiasson et al., 2022). Additional studies using tasks such as the stop-signal task and emotional conflict task have revealed altered activation in regions involved in inhibitory control and emotional processing (Stinson et al., 2024; Marusak et al., 2015). The absence of significant findings in our study may be attributed to our younger sample (ages 9–11), the use of resting-state rather than task-based fMRI, or effects occurring in brain regions or networks not included in our current analyses.

### Limitations and Future Directions

Our findings align with prior evidence suggesting that the neurodevelopmental impact of childhood maltreatment may vary by sex, subtype of adversity, and developmental timing (Teicher & Samson, 2016; Damme et al., 2024). Although research has identified changes across a wide range of brain regions — including the hippocampus, amygdala, PFC, OFC, ACC, and large-scale networks such as SN, DMN, FPN, and CON (Teicher & Samson, 2016; Bick & Nelson, 2016; van Harmelen et al., 2010; Hanson et al., 2010; van Oort et al., 2025; Chahal et al., 2022; Yu et al., 2019) — results have been mixed and sometimes contradictory, depending on the developmental stage at which imaging is conducted, the age at which maltreatment occurred, and its duration. These inconsistencies may reflect methodological heterogeneity, differences in sensitive periods, or moderating factors such as psychopathology and resilience (McLaughlin et al., 2014; Pechtel et al., 2014; Whittle et al., 2013). Future research using harmonized measures and longitudinal designs is critical to clarifying these complex patterns.

First, the prevalence rates of ACE subtypes in our ABCD sample were relatively low, which may limit the generalizability of our findings to youth with more severe or chronic adversity. Several factors may explain this pattern. (1) Certain ACE subtypes, such as sexual abuse, are more likely to occur after the age range captured in our sample (9–10 years old). For instance, Finkelhor et al. (2014) found that approximately 66% of sexual abuse cases occur between the ages of 12 and 17. (2) The assessment method may have contributed to underreporting. ACE questions were answered either by children or parents; however, some children at this developmental stage may lack the cognitive or emotional maturity to fully comprehend or disclose such experiences, while parents may be reluctant to report abuse-related events due to stigma or fear of consequences. Notably, abuse-related questions were answered only by parents and showed higher rates of missing data, potentially reflecting social desirability bias. (3) Our ACE measures were binary and categorized by subtype, rather than using cumulative ACE scores (e.g., 0–21 or categories such as 0 [none], 1–3 [moderate], and 4+ [high]) or dimensional models (e.g., threat vs. deprivation) (e.g., Damme et al., 2024; Stinson et al., 2024; Xiao et al., 2024), which may have reduced sensitivity to a full range of adversity severity.

Second, we did not consider the overlapping of ACE subtypes. Prior studies have shown substantial co-occurrence among ACE subtypes and overlapping types of maltreatment may compound risk (Dong et al., 2004; Finkelhor et al., 2014; McLaughlin et al., 2014), so we examined the overlap between subtypes in our sample and found it to be relatively low. For example, the largest number of participants who experience more than one ACE subtype is 17 (experiencing both physical neglect and emotional neglect). This may reflect the younger age of participants (9–10 years), where maltreatment might not fully happen, underreporting, or the binary categorization of ACEs used in our study. Nonetheless, the low overlap limited our ability to investigate the interactive or cumulative effects of multiple ACE exposures, which may be important for understanding more complex neurodevelopmental outcomes.

Despite these limitations, the prevalence rates and low subtype overlap observed in our study still reflect real-world exposure patterns in a large, demographically diverse, community-based sample. Prior pediatric neuroimaging studies often rely on small, high-risk samples ranging from 38 to 225 participants (De Bellis et al., 2002; Carrion et al., 2001; Hanson et al., 2010; De Brito et al., 2013; Chahal et al., 2022). In contrast, our study leverages a much larger, population-based sample, enabling the detection of subtler associations between early adversity and neurodevelopment and better capturing the variability in ACE exposure as it naturally occurs in the general population. Future research should broaden and refine the characterization of childhood adversity. This includes distinguishing between ACE subtypes (e.g., parental incarceration, household mental illness, or divorce), exploring their co-occurrence and interactive effects, and examining whether specific combinations exert unique effects on neurodevelopment. Greater attention should be paid to sex differences, particularly during transitional periods such as adolescence, where girls appear disproportionately vulnerable to later psychopathology following ACE exposure (Whitaker et al., 2021). Future research should also consider incorporating child self-report tools, such as the PhenX Adverse Life Events module and the Childhood Trauma Questionnaire (CTQ), to capture children’s subjective experiences and provide a more nuanced, developmentally appropriate assessment of adversity, including their perceptions, cognitive appraisal, and potential coping strategies during maltreatment.

Third, our analyses relied solely on cross-sectional baseline data, with participants scanned at ages 9–10, limiting our ability to conclude the directionality or progression of brain changes to ACEs. Fourth, the ABCD dataset lacks important contextual factors related to adverse experiences, including the chronicity of abuse/neglect (i.e., age of onset, duration, frequency) and perpetrator-victim relationship—factors that may significantly moderate neural outcomes (Tomoda et al., 2009; Ketring & Feinauer, 1999; Luby et al., 2013). Fifth, although we did not control for psychopathology (e.g., CBCL scores), this decision was intentional. Our conceptual framework views ACEs and psychopathology as intertwined, rather than as independent contributors to brain development. Sixth, we did not correct for possibly related factors such as pubertal status and BMI. Nonetheless, future work should explicitly model psychopathology to examine which children exposed to adversity remain resilient, and what neural, psychological (e.g., coping strategies, self-esteem; Suzuki & Tomoda, 2015), or social factors (e.g., parenting quality, socioeconomic adversity; Cao et al., 2024; Luby et al., 2013) contribute to that resilience. For instance, increased fractional anisotropy in the corpus callosum has been linked to resilience among adolescents facing high stress exposure (Teicher & Samson, 2016). Functional brain changes may also provide compensatory support: studies suggest that increased connectivity, particularly in females or in those exposed to threat-related ACEs, may buffer against negative outcomes (Brieant et al., 2021; Xiao et al., 2024). A clearer understanding of these protective mechanisms could inform interventions aimed at fostering resilience in at-risk youth.

Given our findings, the next step might be to explore whether hippocampal changes in sexually abused boys and lOFC alterations in emotionally abused boys predict later psychopathology. Longitudinal data with multi-modal imaging will be critical to assessing whether functional connectivity changes mitigate structural deficits and whether connectome-based patterns might serve as predictive markers of resilience. Transdiagnostic approaches will be essential for disentangling the complex, dynamic pathways linking early adversity, brain development, and long-term health. This would advance the goals of precision medicine and help define phenotypes that are more responsive to preventive and therapeutic interventions.

In summary, this study examined the relationship between adverse childhood experiences and brain structural changes in adolescents, highlighting notable sex differences. The results suggest that boys who experience sexual abuse show greater hippocampal volume reduction, while those who experience emotional abuse exhibit more lOFC volume reduction, compared to girls. Although boys are generally less likely to be exposed to sexual abuse than girls, their brain structures may be more vulnerable to certain types of adversity. This study is a first step toward a more comprehensive understanding of how early adversity affects brain development. Future longitudinal work is needed to establish causality and inform effective interventions.

## Data Availability

The data used in this study are publicly available from the Adolescent Brain Cognitive Development (ABCD) Study through the National Institute of Mental Health Data Archive (NDA) at https://nda.nih.gov/abcd. Access requires data use certification.

